# Investigating childhood maltreatment as a modifier of genetic risk for cardiovascular disease in the UK Biobank

**DOI:** 10.1101/2022.01.26.22269809

**Authors:** Helena Urquijo, Ana Gonçalves Soares, Abigail Fraser, Laura D Howe, Alice R Carter

**Author notes:** **Corresponding Author:** Helena Urquijo, MRC Integrative Epidemiology Unit, University of Bristol, Oakfield House, Oakfield Grove, BS8 2BN, Bristol, United Kingdom.

## Abstract

Cardiovascular disease (CVD) is influenced by genetic and environmental factors. Childhood maltreatment is associated with CVD and may modify genetic susceptibility to cardiovascular risk factors. We used genetic and phenotypic data from 100,833 White British UK Biobank participants (57% female; mean age = 55.9 years). We regressed nine cardiovascular risk factors/diseases (alcohol consumption, body mass index [BMI], low-density lipoprotein cholesterol, lifetime smoking behaviour, systolic blood pressure, atrial fibrillation, coronary heart disease, type 2 diabetes, and stroke) on their respective polygenic scores (PGS) and self-reported exposure to childhood maltreatment. Effect modification was tested on the additive and multiplicative scales by including a product term (PGS*maltreatment) in regression models.

On the additive scale, childhood maltreatment accentuated the effect of genetic susceptibility to higher BMI. Individuals not exposed to childhood maltreatment had an increase in BMI of 0.12 SD (95% CI: 0.11, 0.13) per SD increase in BMI PGS, compared to 0.17 SD (95% CI: 0.14, 0.19) in those exposed to all types of childhood maltreatment. On the multiplicative scale, similar results were obtained for BMI though these did not withstand to Bonferroni correction. There was little evidence of effect modification by childhood maltreatment in relation to other outcomes, or of sex-specific effect modification.

Our study suggests the effects of genetic susceptibility to a higher BMI may be moderately accentuated in individuals exposed to childhood maltreatment. However, gene*environment interactions are likely not a major contributor to the excess CVD burden experienced by childhood maltreatment victims.

## Introduction

Cardiovascular disease (CVD) is the leading cause of death worldwide.^1^ Many CVD subtypes have complex aetiologies, affected by both genetic and environmental factors.^2,3^ This suggests that genetic and environmental factors may interact with each other and modify susceptibility to disease. Childhood maltreatment increases the risk of a number of CVD subtypes and related diseases, including coronary heart disease (CHD), stroke and type 2 diabetes mellitus (T2DM).^4,5^ Evidence also suggests there is a dose-response relationship between childhood maltreatment and CVD, with disease risk increasing according to the number of types of maltreatment experienced by an individual.^6–13^ While this growing body of evidence indicates childhood maltreatment is associated with higher CVD risk, it is unknown whether this risk is disproportionate in genetically susceptible individuals.

Single nucleotide polymorphisms (SNPs) detected in genome-wide association studies (GWAS) have been used to construct polygenic scores (PGSs) for a trait or disease. These estimate the genetic risk carried by an individual by adding the effects of all relevant variants in the genome. The polygenic nature of many diseases, including CVD, suggests these scores may be more informative when investigating interactions or effect modification by environmental factors, in comparison to using a single associated gene.^14^ PGSs have previously been employed to assess gene*environment interactions or effect modifications for cardiovascular traits, including for educational achievement, neighbourhood and obesogenic environments.^15–17^ Identifying these effects may help to understand the relationship between early life environmental exposures and CVD risk in adult life.

This study aims to investigate whether childhood maltreatment modifies genetic susceptibility to a range of cardiovascular risk factors and CVD subtypes, using data from the UK Biobank cohort. Where two risk factors are associated with the outcome, evidence of effect modification is expected on either the additive or the multiplicative scale, or on both scales.^18,19^ Therefore, in this analysis we investigate both scales. Of greatest relevance to public health is the direction and magnitude of any interaction. We hypothesise that childhood maltreatment accentuates the effect of polygenic predisposition to CVD subtypes and risk factors. We also investigate whether any potential effect modification differs according to sex, as some studies have detected differences in the association between childhood maltreatment and cardiovascular risk^13,20–23^

## Methods

### UK Biobank

UK Biobank recruited 503,317 adults aged 37-73 years at baseline, between 2006 and 2010.^24^ At baseline, data was collected on health and lifestyle information, physical measurements and biological samples. Some follow-up questionnaires and clinics have taken place, including an online mental health questionnaire in 2016, which asked about childhood maltreatment. This was sent to participants who provided an email address (N=339,092) and 157,366 (46.4%) responses were obtained. This analysis includes 100,833 participants of White British ancestry who completed questions on childhood maltreatment and had genetic information. Some individuals were missing CVD risk factor information and thus sample sizes for analyses ranged from n=93,001 to n=100,833 (Table 1).

**Table 1:** Descriptive statistics of study sample and entire Biobank sample (excluding withdrawn participants). For maltreatment types, exposure is determined according to cut-off criteria described in Table S1.

| Variable | Analysis sample N = 100833 |  |  | Full UK Biobank N = 502 493 |  |  |
| --- | --- | --- | --- | --- | --- | --- |
| Continuous | N | Mean (SD) |  | N | Mean (SD) |  |
| Age | 100833 | 55.88 (7.67) |  | 502493 | 56.53 (8.10) |  |
| BMI | 100659 | 26.57 (4.38) |  | 499389 | 27.43 (4.80) |  |
| Drinks per week | 100529 | 8.38 (8.66) |  | 498251 | 7.79 (9.05) |  |
| LDL cholesterol | 95809 | 3.64 (0.82) |  | 468696 | 3.56 (0.87) |  |
| Lifetime smoking | 95845 | 0.25 (0.55) |  | 318071 | 0.34 (0.67) |  |
| Systolic blood pressure | 93001 | 136.37 (18.08) |  | 456954 | 137.78 (18.63) |  |
| Number of siblings | 99629 | 1.78 (1.47) |  | 492858 | 2.23 (2.00) |  |
| Categorical | N | Frequency (%) |  | N | Frequency (%) |  |
| Sex | 100833 | Female | 57 641 (57) | 502493 | Female | 273 378 (54) |
| Maternal smoking | 88907 | No | 63 353 (71) | 434357 | No | 307 274 (71) |
| Atrial fibrillation (incident) | 100833 | Control | 98 735 (97) | 496107 | Control | 480 339 (97) |
|  |  | Case | 2 098 (2) |  | Case | 15 768 (3) |
| Type 2 Diabetes (incident) | 100833 | Control | 99 449 (99) | 493064 | Control | 47 2429 (96) |
|  |  | Case | 1 384 (1) |  | Case | 20 635 (4) |
| Coronary Heart Disease (incident) | 100833 | Control | 97 801 (97) | 481868 | Control | 45 9019 (95) |
|  |  | Case | 3 032 (3) |  | Case | 22 849 (5) |
| Stroke (incident) | 100833 | Control | 99 838 (99) | 497488 | Control | 48 7421 (98) |
|  |  | Case | 995 (1) |  | Case | 10 067 (2) |
| Physical abuse | 100833 | Yes | 17 530 (17) | 156982 | Yes | 29 793 (19) |
| Emotional abuse |  |  | 14 535 (14) | 156886 |  | 24 544 (16) |
| Sexual abuse |  |  | 8 191 (8) | 155488 |  | 13 646 (9) |
| Physical neglect |  |  | 15 286 (15) | 156266 |  | 25 723 (16) |
| Emotional neglect |  |  | 21 189 (21) | 156712 |  | 35 290 (23) |
| Childhood maltreatment score | 100833 | 0 | 55 910 (55) | 153625 | 0 | 82 398 (54) |
|  |  | 1 | 25 570 (25) |  | 1 | 39 207 (26) |
|  |  | 2 | 10 846 (11) |  | 2 | 17 482 (11) |
|  |  | 3 | 5 299 (5) |  | 3 | 8 801 (6) |
|  |  | 4 | 2 468 (2) |  | 4 | 4 390 (3) |
|  |  | 5 | 740 (1) |  | 5 | 1 347 (1) |

### Childhood maltreatment

The types of maltreatment assessed comprised of emotional neglect, physical neglect, emotional abuse, physical abuse and sexual abuse.^25,26^ A summary score of childhood maltreatment was calculated as per the Childhood Trauma Screener,^25^ reflecting the cumulative number of maltreatment types experienced by an individual, spanning from 0 to 5 (Table S1).

### Cardiovascular risk factors and diseases

This study included 3 CVD subtypes and 6 risk factors for CVD. Risk factors were considered if there is evidence for a causal association with CVD from randomised control trials (RCT) or Mendelian randomisation studies (see Table S2) - a statistical approach using randomly allocated genetic variants as instrumental variables ^27^ - and have GWAS information with summary statistics available.

Quantitative traits (body mass index (BMI), alcohol consumption measured as drinks per week, low-density lipoprotein cholesterol (LDL-C), lifetime smoking behaviour and systolic blood pressure (SBP) were determined at baseline through physical or biological measurements, or self-reported for alcohol and smoking behaviours.

Incidence of CVD subtypes (atrial fibrillation (AF), CHD and stroke) and of risk factor T2DM were determined through linked mortality data, hospital episode statistics and Scottish morbidity and mortality records (SMR), defined using ICD-9 and ICD-10 codes (Table S3).

The period of follow-up occurred between the date of baseline assessment (between 2006 and 2010) and the most recent available linked hospital inpatient data, from May 2017.

### Polygenic scores

Summary statistics were obtained from the most recent GWAS (Table 2). Where the most recent GWAS included UK Biobank participants, the next most suitable study was used to avoid estimate inflation through sample overlap. PGSs were calculated by multiplying the number of effect alleles by the relative effect estimate of the SNP on the phenotype and summing together the alleles.

**Table 2:** Summary characteristics for each GWAS used to derive external weights in polygenic scores.

| Phenotype | Author/consortium | Population | Sample Size (cases) | Unit |
| --- | --- | --- | --- | --- |
| Alcohol consumption | GWAS and Sequencing Consortium of Alcohol and Nicotine use <sup>31</sup> | European (Summary statistics excluding UK Biobank) | 630 154 | Drinks per week |
| Body mass index | Genetic Investigation of Anthropometric Traits <sup>48</sup> | European ancestry | 339 224 | SD (kg/m<sup>2</sup>) |
| Low-density lipoprotein cholesterol | Global Lipids Genetics consortium <sup>49</sup> | European ancestry | 188 578 | SD (circulating lipids) |
| Smoking | Wootton *et al* <sup>50</sup> | White British (split sample GWAS of UK Biobank, see Supplementary Methods) | 318 147 | SD (Lifetime smoking index) |
| Systolic blood pressure | Carter *et al* <sup>51</sup> | White British (split sample GWAS of UK Biobank, see Supplementary Methods) | 318 147 | SD (mm/Hg) |
| Atrial fibrillation | Ellinor *et al* <sup>52</sup> | European ancestry | 59 133 (6 707) | Log odds ratio |
| Coronary heart disease | CARDIoGRAMplusC4D <sup>53</sup> | Predominantly European (77%) | 184 305 (60 801) | Log odds ratio |
| Type 2 diabetes | DIAbetes Genetics Replication And Meta-analysis <sup>54</sup> | European ancestry | 159 208 (26 276) | Log odds ratio |
| Stroke | MEGASTROKE <sup>55</sup> | Predominantly European (85%) | 521 612 (67 162) | Log odds ratio |

PGSs were generated at P value thresholds of <5×10^−8^ (genome-wide significance), <0.05 and <0.5. The greater the P value threshold for SNPs, the greater the variance explained by the score (Table S4), but the greater the chance of capturing pleiotropic effects. Resulting PGSs were standardised, reflecting a one standard deviation increase in PGS.

Full details of the derivation of PGSs, GWAS studies used and generation of phenotypic measures can be found in the Supplement.

### Exclusion criteria

Quality control of UK Biobank genetic data was performed in accordance with the MRC-IEU Quality Control pipeline, as previously described.^28^ In brief, participants with sex mismatch (comparing genetic sex with reported sex) and those with sex chromosome aneuploidy were excluded from the study. Participants of White British ancestry were those who self-reported as “White British” and supported by genetic principal component (PC) analysis. Related participants were excluded according to kinship coefficients provided by the UK Biobank until no related pairs remained.^28^

Individuals missing age, sex, genetic data or who did not have complete information for childhood maltreatment were excluded from the sample (Figure 1). Individuals missing risk factor or disease data were excluded from relevant analyses. To prevent the detection of effects caused by reverse causality, participants were excluded if they experienced any diagnosis of AF, T2DM, CHD or stroke prior to baseline. Prevalent cases were ascertained by comparing the date of diagnosis in the linked hospital inpatient data to the date of baseline visit.

**Figure 1:**
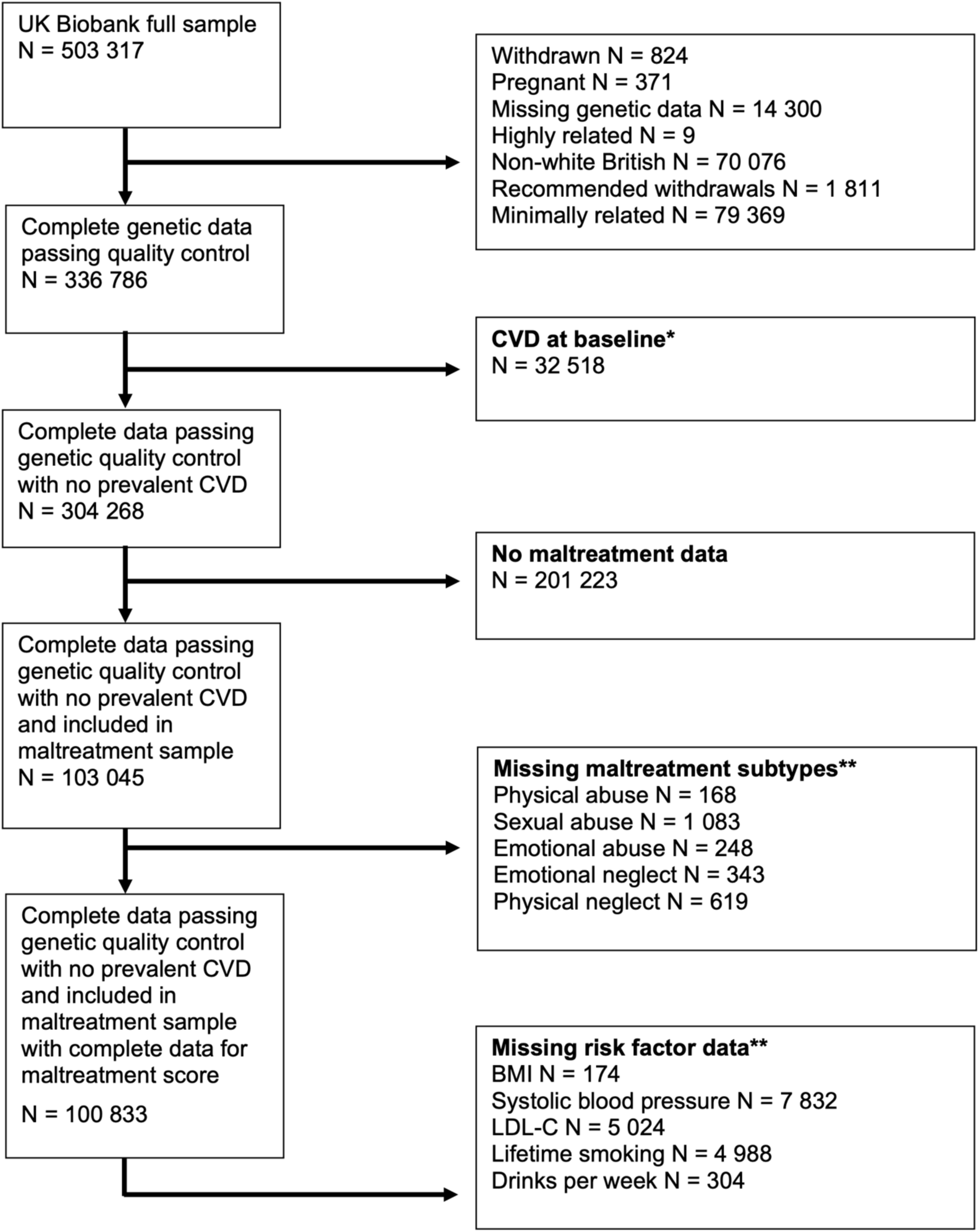
Study exclusion criteria. *CVD = stroke, atrial fibrillation, coronary heart disease or risk factor type 2 diabetes. **Participants can be missing more than one variable, so the total excluded is less than the sum of the missing data of all traits.

### Statistical analysis

Statistical analyses were carried out using Stata16-MP. Analysis code is available at github.com/helenaurquijo/gxe_cvd_maltreatment. Study data used will be archived with UK Biobank and was carried out under project 19278.

#### Association of childhood maltreatment with outcomes

Multivariable linear and logistic regression models, adjusting for sex and age, were used to estimate the association between the childhood maltreatment score and each of the nine risk factors/diseases.

#### Effect modification of childhood maltreatment on genetic susceptibility

For continuous traits, the association of childhood maltreatment score and each PGS with the respective trait was estimated on the additive scale via multivariable linear regression. On the multiplicative scale, the association was estimated via multivariable linear regression for the natural log of the trait. For binary traits, the association of childhood maltreatment score and each PGS with the respective disease trait was estimated on the additive scale via multivariable linear regression. On the multiplicative scale, the association was estimated via multivariable logistic regression. All regressions were adjusted for sex, age and 40 genetic PCs. Continuous phenotypic measures of traits were standardised to reflect a change in SD of trait per 1 SD increase in PGS. Coefficients for binary traits reflect a change in risk per SD increase in PGS.

To test for effect modification of genetic predisposition by exposure to childhood maltreatment, the same regression models were run including a product of the PGS and maltreatment score as an interaction term. Effect modification was evaluated by the size and precision of the coefficient for effect modification, and the P value for the effect modification coefficient considering Bonferroni correction.

To address the risk of type 1 error due to multiple comparisons, a Bonferroni correction was applied by dividing the standard 0.05 P value threshold by the number of outcomes we considered in each analysis (nine), yielding a threshold of P=0.006. The use of this correction approach was conservative as the considered traits were not independent from each other.

#### Secondary analyses

Sex-stratified analyses were carried out, and a 3-way interaction term (i.e. PGS*maltreatment*sex) on the respective risk factor or CVD subtype was included.

Main analyses were repeated using PGSs generated with less stringent P value thresholds at 0.05 and 0.5, as described previously.

Finally, sensitivity analyses were performed by replicating the main analyses with adjustments for covariate interactions (e.g. sex*PGS, age*maltreatment…) to avoid specification error.^14^ Separately, maternal smoking and number of siblings (proxies of childhood socioeconomic position), which may confound associations between childhood maltreatment and CVD, were included in the adjustment.

## Results

### UK Biobank sample

Eligible participants had a mean age at baseline of 56 years (SD: 8.0 years) and 57% were female. Most participants (55%) reported no exposure to any type of childhood maltreatment. The most frequently reported type of maltreatment was emotional neglect (21%) and sexual abuse was the least reported (8%) (Table 1). Our study sample did not differ from the rest of the UK Biobank in terms of exposure to maltreatment, cardiovascular risk and cases of CVD.

With the genome-wide significance threshold (P<5×10^−8^), variance explained by the PGSs ranged between 0.06% (smoking) and 15% (systolic blood pressure) (Table S4).

### Association of childhood maltreatment with cardiovascular risk factors and disease

Childhood maltreatment score was associated with higher BMI (β: 0.07 SD; 95% CI: 0.06, 0.07) and lifetime smoking, as well as with T2DM and CHD incidence (Table S5). However, childhood maltreatment was negatively associated with systolic blood pressure (β: −0.011 SD; 95% CI: −0.017, - 0.005). Childhood maltreatment was positively associated with all other outcomes, but the confidence intervals spanned the null.

### Effect modification by childhood maltreatment

On the additive scale, there was little evidence that childhood maltreatment modified the effect of the PGSs for all risk factors/diseases except for BMI (Table S6 and Figure 2). The effect of genetic susceptibility to higher BMI was accentuated by childhood maltreatment score by 0.009 SD (95% CI: 0.003, 0.014; P_effect modification_: 0.003). This indicates, in those unexposed to childhood maltreatment, each SD increase in the BMI PGS was associated with 0.12 SD higher BMI (95% CI: 0.11, 0.13), whilst in those with a maltreatment score of 5, each SD increase in the PGS was associated with 0.17 SD higher BMI (95% CI: 0.14, 0.19). All other interaction effect estimates were positive, with the exception of SBP, and extremely small.

**Figure 2:**
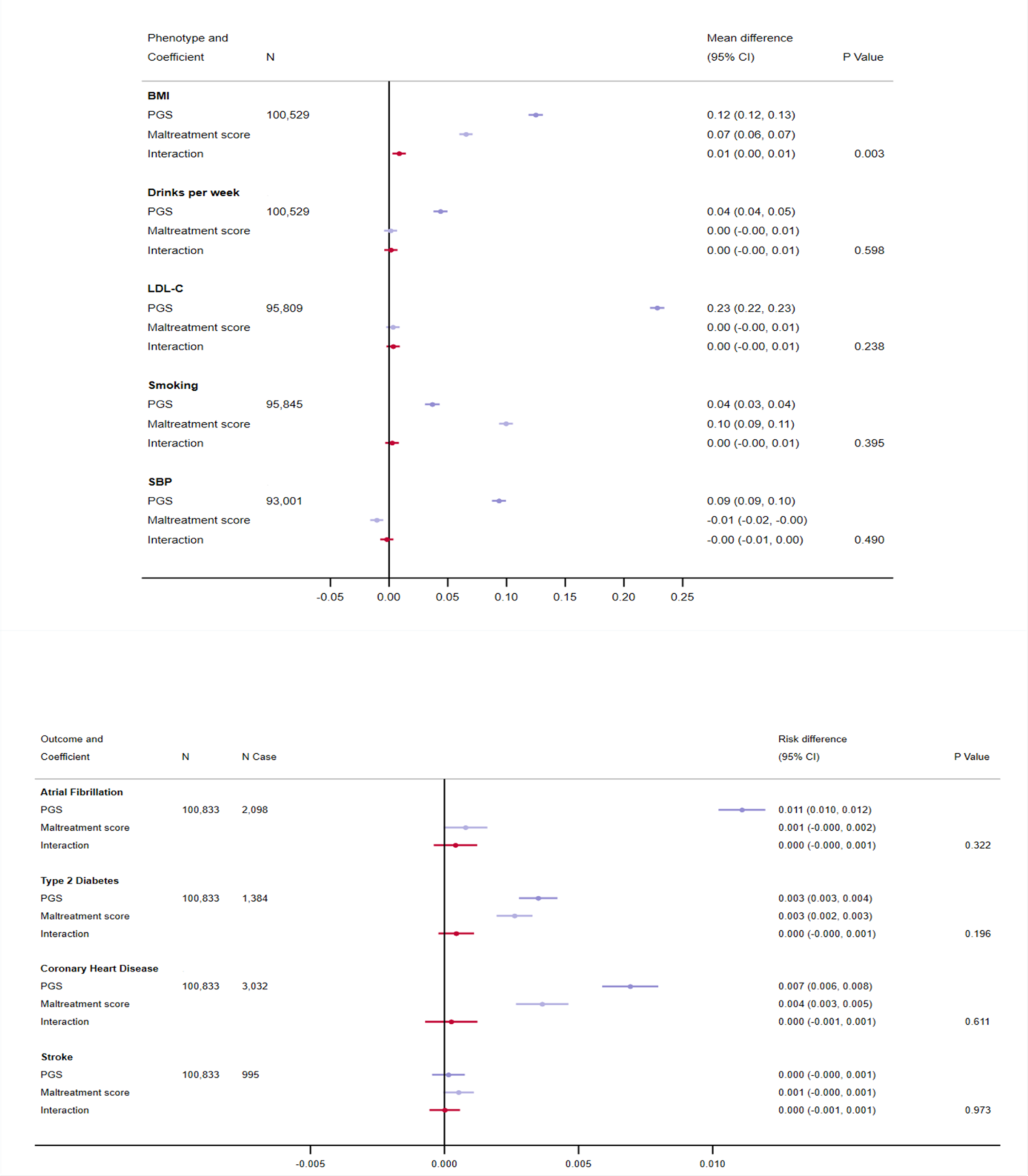
Effect modification by exposure to childhood maltreatment on genetic predisposition to cardiovascular risk factors and disease on the additive scale at P<5×10^−8^. Analyses were adjusted for age, sex and 40 genetic principal components. Traits are grouped according to whether they are binary or continuous. BMI=body mass index; LDL-C=low-density lipoprotein cholesterol; SBP=systolic blood pressure; PGS=polygenic score.

On the multiplicative scale, similar yet less precise results were obtained (Table S7 and Figure 3). The effect of genetic susceptibility to higher log BMI was accentuated by childhood maltreatment, through an increase of 0.007 SD (95% CI 0.001, 0.013) per unit increase in maltreatment score. Nonetheless, the interaction estimate did not withstand Bonferroni correction (P_effect modification_: 0.015). The coefficient for effect modification was in the positive direction for alcohol consumption, LDL-C, smoking, AF and stroke, and in the negative direction for SBP, CHD and T2DM. However, for all outcomes the estimates were close to the null and confidence intervals were narrow.

**Figure 3:**
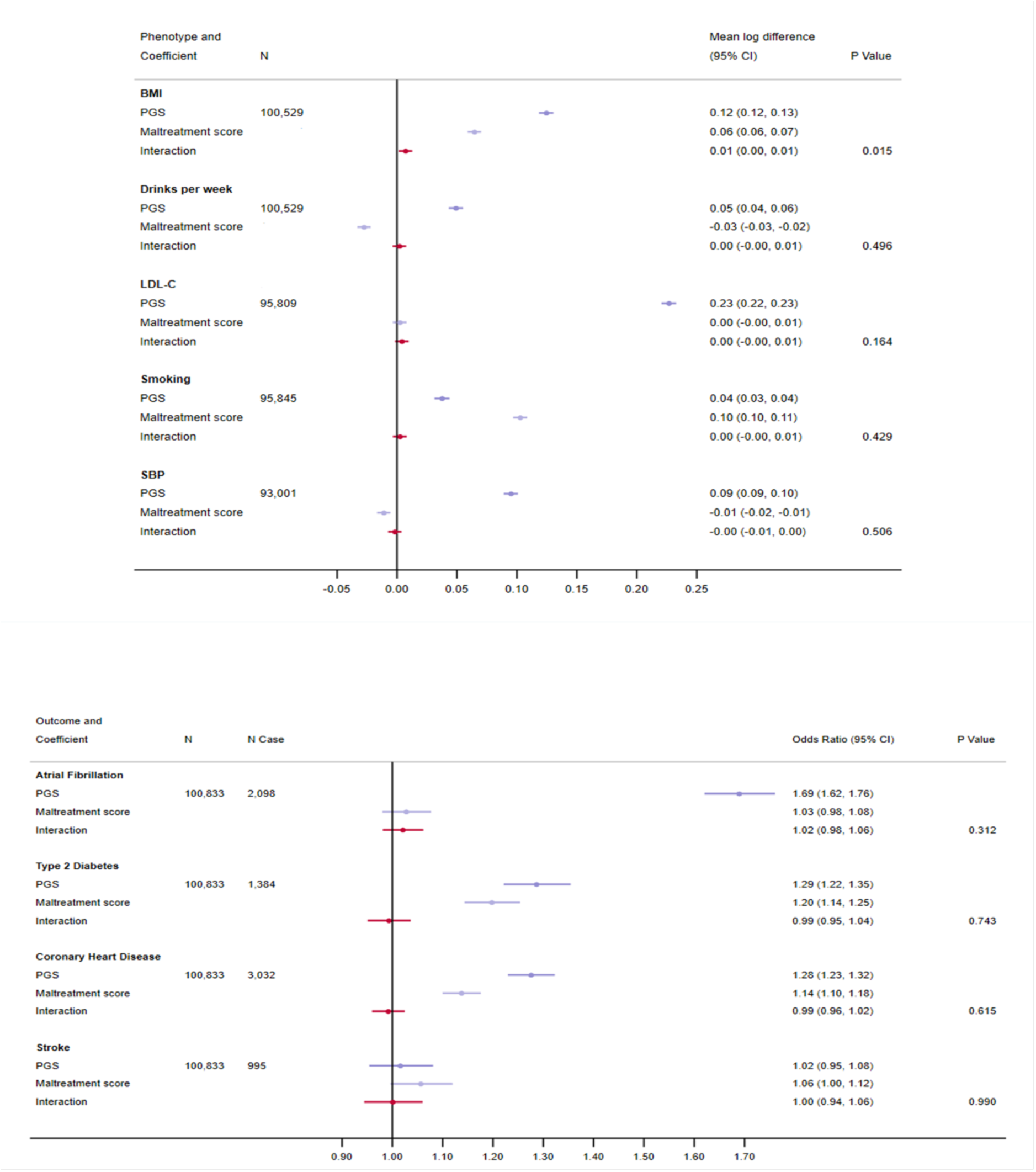
Effect modification by exposure to childhood maltreatment on genetic predisposition to cardiovascular risk factors and disease on the multiplicative scale at P<5×10^−8^. Analyses were adjusted for age, sex and 40 genetic principal components. Traits are grouped according to whether they are binary or continuous. BMI=body mass index; LDL-C=low-density lipoprotein cholesterol; SBP=systolic blood pressure; PGS=polygenic score.

### Secondary analyses

Sex was not found to modify the effects observed in the main analyses on either the additive or the multiplicative scale (Tables S8 and S9).

Analyses repeated with PGSs generated at P value thresholds of P≤0.05 and P≤0.5 found childhood maltreatment score to accentuate the effect of genetic susceptibility to higher BMI and greater lifetime smoking, on both the additive and multiplicative scale (Tables S10 and S11).

Analyses adjusting for covariate (age, sex and genetic PCs) interactions with childhood maltreatment score and PGSs yielded similar results, though the interaction coefficients for BMI became less precise, decreasing the strength of evidence for effect modification on the multiplicative scale (Table S12).

Analyses adjusting for maternal smoking and number of siblings yielded similar results to main analyses (Table S13).

## Discussion

We found evidence that childhood maltreatment accentuates the effect of polygenic susceptibility to BMI on observed BMI but no other cardiovascular risk factors or outcomes considered. Effects were consistent across the additive and multiplicative scales, though evidence of effect modification for BMI was weaker on the multiplicative scale. Additionally, we found no evidence of sex differences in these effects.

### Results in context

Few studies have investigated whether childhood maltreatment interacts with, or modifies the effect of genetic susceptibility to a range of cardiovascular risk factors/outcomes. In a candidate gene approach, Gooding and colleagues found no association between childhood maltreatment and hypertension in young adulthood, nor the presence of effect modification by the SLC64A genotype.^29^ In our study of adults, we found a negative association between childhood maltreatment and SBP. In a study with 161 individuals with major depression, Opel and colleagues also found childhood maltreatment modified polygenic susceptibility to BMI but in the opposite direction, with exposure to maltreatment reducing the effect of polygenic susceptibility on BMI.^30^ These opposing findings could be due to different definitions of maltreatment (binary versus continuous score), or different study populations (depressive patients versus population cohort).

We expected to detect effect modification for all outcomes given (a) the association between PGSs and respective phenotypes and (b) the positive associations between childhood maltreatment and CVD subtypes present in our study sample (see Table S4 and interpreting interactions section below). It is difficult to determine whether the lack of effect modification for most outcomes is truly null, underpowered, or due to other factors such as the definition used for risk factors. We have used drinks per week to measure alcohol intake, while previous studies have used units of alcohol per week.^31^ For drinking and smoking, misclassification bias may occur, which may be differential misclassification if this differs between participants with and without exposure to maltreatment. Integrating the five types of childhood maltreatment into a score, while relevant in the light of dose-response relationships between childhood maltreatment and CVD, may miss maltreatment type-specific effects.

Reports of sex differences in the association between childhood maltreatment subtypes and CVD have been mixed,^6,13,32–35^ however we did not identify any sex-specific differences. Replication and validation using other cohorts may elucidate whether we missed modification and sex-specific effects due to lack of statistical power.

Positive associations between childhood maltreatment and CVD subtypes and BMI have been reported extensively ^5,36^ but associations with other CVD risk factors such as blood pressure and lipids are less established.^36^ We replicated previously observed associations between maltreatment increasing BMI ^32–34^, smoking behaviour ^6,37^, and decreasing SBP.^38,39^

### Interpretability of interactions and effect modifications

Interaction and effect modification are terms commonly used interchangeably despite representing subtly different concepts. The underlying statistical model is identical.^40^

In an interaction, the effects of two causal and independent risk factors are studied, to assess whether their joint exposure leads to a different outcome compared with their independent associations. In effect modification, the effect of one exposure differs (or is modified) according to the value of a second exposure. These exposures are not necessarily causal.^18^ Both interaction and effect modification can be estimated on the additive or the multiplicative scale. If two exposures are associated with the outcome, an interaction should be detected on at least one of the scales given sufficient statistical power.

In the case of childhood maltreatment, although consistent associations with CVD exist, causality is difficult to assess and therefore the term effect modification better reflects the model considered here. We investigated whether the effect of polygenic susceptibility to a certain trait differed according to values of the childhood maltreatment score. Previous studies have typically only tested the additive scale for interaction.^15,16,19,30^ Because effect modification can occur on either the additive scale or the multiplicative scale, we analysed both scales.^18,19^

### Relevance for public health

The size of the interaction terms of the few effect modifications detected in this study were very small, even for models employing PGSs with SNPs included using a less stringent significance thresholds. This implies that the clinical impact of these effect modifications is very low, suggesting gene*environment effect modifications are not a major contributor to the excess burden of cardiovascular disease experienced by people who have experienced child maltreatment.

### Strengths and limitations

The major strength of our study is the use of a single large cohort, the UK Biobank, giving a large sample size and homogenous measures for our variables. While many studies rely on self-reported measures of phenotypes or disease outcomes, we used hospital and mortality records for CVD and calibrated physical measurements to define phenotypes. Reverse causality can bias analyses, whereby the diagnosis of CVD may lead to medical treatment or altered lifestyle behaviours altering the relative importance of the genotype on disease risk. We therefore excluded all participants with prevalent cases of CVD.

Another strength of this study lies in the use of PGSs rather than a single candidate gene approach, where candidate gene interactions have been shown to be spurious.^41^ We also repeated analyses using PGSs generated at different significance thresholds, as PGSs including variants detected at less stringent P value thresholds may be more predictive of disease.^42^ Finally, PGSs are not subject to the same confounding as environmental exposures because they are determined at conception, though they may be affected by population structure. All models were adjusted for all 40 genetic PCs calculated by the UK Biobank to mitigate this bias.

A number of limitations remain. Participation in the mental health questionnaire, an inclusion criterion for our study, has been associated with lower BMI, higher socioeconomic position, and having no T2DM or depression, compared to the rest of the UK Biobank cohort.^43^ Estimates may be biased by selection bias, where UK Biobank participants are more likely to be female, of higher socioeconomic position, less likely to be obese, to smoke, to drink alcohol on a frequent basis and to experience CVD compared to the rest of the population.^24^

Childhood maltreatment was determined using self-reported retrospective measures, which could introduce recall bias. An alternative approach would be verifying self-reports with records from social services or other child protecting entities. However, only a fraction of maltreatment cases are reported ^44^ and thus many cases might be missed. Another limitation is that despite the adjustment for maternal smoking and number of siblings, these might not be the most comprehensive indicators of childhood SEP. Maternal smoking may not be a robust indicator of SEP for this study’s participants as the social patterning of maternal structure was different in the time these participants were born in comparison to current day. This means residual confounding, by childhood SEP or other unmeasured sources, may have biased our findings.

We applied a Bonferroni correction when considering the effect modification P values to address the risk of type 1 error due to multiple tests. This may be overly conservative as cardiovascular risk factors/diseases are correlated phenotypes. However, considering a less stringent P value threshold of P<0.05, there was still little evidence of an interaction between childhood maltreatment and any outcome other than BMI.

Finally, there are caveats with PGSs. Residual confounding by population structure may remain even after adjusting for 40 genetic PCs ^45^. While our study only included participants of White British Ancestry and used summary statistics from similar populations, some of the PGSs explained a low proportion of the trait’s variance. More so, our findings have limited generalizability to non-White European populations, as PGSs have been found to have very limited portability across ancestries ^46^.

The disproportionate scrutinising of a single ancestral background is an impediment in our understanding of CVD aetiology and the clinical application of PGSs. Indeed, 83% of GWASs released between 2005 and 2018 were on individuals of White European Ancestry ^47^, underscoring the importance of the need for trans-ethnic GWASs and the replication of studies such as ours in other populations.

## Conclusion

In this analysis on White British individuals from the UK Biobank, we exposure to childhood maltreatment can moderately exacerbate the effects of a genetic propensity to higher BMI. However, we found little evidence that childhood maltreatment modified the effects of other cardiovascular polygenic scores. Our results suggest gene*environment interactions are not a major contributor to the excess burden of CVD experienced by people who have experienced child maltreatment.

## Data Availability

All data produced in the present work are contained in the manuscript and in supplementary file. The code used in this analysis is available at github.com/helenaurquijo/gxe_cvd_maltreatment.

## Acknowledgements

Quality Control filtering of the UK Biobank data was conducted by R.Mitchell, G.Hemani, T.Dudding, L.Corbin, S.Harrison, L.Paternoster as described in the published protocol (doi: 10.5523/bris.1ovaau5sxunp2cv8rcy88688v). The MRC IEU UK Biobank GWAS pipeline was developed by B.Elsworth, R.Mitchell, C.Raistrick, L.Paternoster, G.Hemani, T.Gaunt (doi: 10.5523/bris.pnoat8cxo0u52p6ynfaekeigi).

## Author contributions

HU cleaned and analysed the data, interpreted the results, wrote and revised the manuscript. ALGS and ARC had access to the data and assisted in data preparation. ALGS, AF, LDH and ARC all designed the study, interpreted the results, critically reviewed and revised the manuscript and provided supervision for the project. HU and ARC serve as guarantors of the paper. The corresponding author attests that all listed authors meet authorship criteria and that no others meeting the criteria have been omitted.

## Abbreviations and Acronyms

AF: Atrial fibrillation
BMI: Body mass index
CHD: Coronary heart disease
CVD: Cardiovascular disease
GWAS: Genome-wide association studies
LDL-C: Low-density lipoprotein cholesterol
PC: Principal components
PGS: Polygenic score
RCT: Randomised control trials
SBP: Systolic blood pressure
SEP: Socioeconomic position
SNP: Single nucleotide polymorphism
T2DM: Type-2 diabetes mellitus

## References

1. Wang H, Naghavi M, Allen C, et al. Global, regional, and national life expectancy, all-cause mortality, and cause-specific mortality for 249 causes of death, 1980-2015: a systematic analysis for the Global Burden of Disease Study 2015. Lancet. 2016;388(10053):1459–1544. doi:10.1016/S0140-6736(16)31012-1

2. Delles C, McBride MW, Padmanabhan S, Dominiczak AF. The genetics of cardiovascular disease. Trends Endocrinol Metab. 2008;19(9):309–316. doi:10.1016/j.tem.2008.07.010

3. Bhatnagar A. Environmental Determinants of Cardiovascular Disease. Circ Res. 2017;121(2):162–180. doi:10.1161/CIRCRESAHA.117.306458

4. Suglia SF, Koenen KC, Boynton-Jarrett R, et al. Childhood and Adolescent Adversity and Cardiometabolic Outcomes: A Scientific Statement from the American Heart Association. Circulation. 2018;137(5):e15–e28. doi:10.1161/CIR.0000000000000536

5. Basu A, McLaughlin KA, Misra S, Koenen KC. Childhood Maltreatment and Health Impact: The Examples of Cardiovascular Disease and Type 2 Diabetes Mellitus in Adults. Clin Psychol Sci Pract. 2017;24(2):125–139. doi:10.1111/cpsp.12191

6. Dong M, Giles WH, Felitti VJ, et al. Insights into causal pathways for ischemic heart disease: Adverse childhood experiences study. Circulation. 2004;110(13):1761–1766. doi:10.1161/01.CIR.0000143074.54995.7F

7. Felitti VJ, Anda RF, Nordenberg D, et al. Relationship of Childhood Abuse and Household Dysfunction to Many of the Leading Causes of Death in Adults. Am J Prev Med. 1998;14(4):245–258. doi:10.1016/S0749-3797(98)00017-8

8. Campbell JA, Walker RJ, Egede LE. Associations Between Adverse Childhood Experiences, High-Risk Behaviors, and Morbidity in Adulthood. Am J Prev Med. 2016;50(3):344–352. doi:10.1016/j.amepre.2015.07.022

9. Scott KM, Von Korff M, Angermeyer MC, et al. Association of childhood adversities and early-onset mental disorders with adult-onset chronic physical conditions. Arch Gen Psychiatry. 2011;68(8):838–844. doi:10.1001/archgenpsychiatry.2011.77

10. Wilson RS, Boyle PA, Levine SR, et al. Emotional neglect in childhood and cerebral infarction in older age. Neurology. 2012;79(15):1534–1539. doi:10.1212/WNL.0b013e31826e25bd

11. Friedman EM, Montez JK, Sheehan CM, Guenewald TL, Seeman TE. Childhood Adversities and Adult Cardiometabolic Health. J Aging Health. 2015;27(8):1311–1338. doi:10.1177/0898264315580122

12. Ho FK, Celis-Morales C, Celis-Morales C, et al. Child maltreatment and cardiovascular disease: Quantifying mediation pathways using UK Biobank. BMC Med. 2020;18(1):1–10. doi:10.1186/s12916-020-01603-z

13. Soares ALG, Hammerton G, Howe LD, Rich-Edwards J, Halligan S, Fraser A. Sex differences in the association between childhood maltreatment and cardiovascular disease in the UK Biobank. Heart. Published online 2020:1310-1316. doi:10.1136/heartjnl-2019-316320

14. Domingue BW, Trejo S, Armstrong-Carter E, Tucker-Drob EM. Interactions between polygenic scores and environments: Methodological and conceptual challenges. Sociol Sci. 2020;7:465–486. doi:10.15195/V7.A19

15. Tyrrell J, Wood AR, Ames RM, et al. Gene–obesogenic environment interactions in the UK Biobank study. Int J Epidemiol. 2017;46(2):559–575. doi:10.1093/ije/dyw337

16. Mason KE, Palla L, Pearce N, Phelan J, Cummins S. Genetic risk of obesity as a modifier of associations between neighbourhood environment and body mass index: an observational study of 335 046 UK Biobank participants. BMJ Nutr Prev Heal. Published online October 5, 2020:bmjnph-2020-000107. doi:10.1136/bmjnph-2020-000107

17. Carter AR. Strengthening causal inference in educational inequalities in cardiovascular disease. Published online 2021.

18. VanderWeele TJ. On the Distinction Between Interaction and Effect Modification. Epidemiology. 2009;20(6):863–871. doi:10.1097/EDE.0b013e3181ba333c

19. Van Der Weele TJ, Knol MJ. A tutorial on interaction. Epidemiol Method. 2014;3(1):33–72. doi:10.1515/em-2013-0005

20. Goodwin RD, Stein MB. Association between childhood trauma and physical disorders among adults in the United States. Psychol Med. 2004;34(3):509–520. doi:10.1017/S003329170300134X

21. Widom CS, Czaja SJ, Bentley T, Johnson MS. A Prospective Investigation of Physical Health Outcomes in Abused and Neglected Children: New Findings From a 30-Year Follow-Up. Am J Public Health. 2012;102(6):1135–1144. doi:10.2105/AJPH.2011.300636

22. Fuller-Thomson E, Bejan R, Hunter JT, Grundland T, Brennenstuhl S. The link between childhood sexual abuse and myocardial infarction in a population-based study. Child Abuse Negl. 2012;36(9):656–665. doi:10.1016/j.chiabu.2012.06.001

23. Duncan AE, Auslander WF, Bucholz KK, Hudson DL, Stein RI, White NH. Relationship Between Abuse and Neglect in Childhood and Diabetes in Adulthood: Differential Effects By Sex, National Longitudinal Study of Adolescent Health. Prev Chronic Dis. 2015;12:140434. doi:10.5888/pcd12.140434

24. Fry A, Littlejohns TJ, Sudlow C, et al. Comparison of Sociodemographic and Health-Related Characteristics of UK Biobank Participants with Those of the General Population. Am J Epidemiol. 2017;186(9):1026–1034. doi:10.1093/aje/kwx246

25. Glaesmer H, Schulz A, Häuser W, Freyberger H, Brähler E, Grabe H-J. Der Childhood Trauma Screener (CTS) - Development and Validation of Cut-Off-Scores for Classificatory Diagnostics. Psychiatr Prax. 2013;40(04):220–226. doi:10.1055/s-0033-1343116

26. Davis KAS, Coleman JRI, Adams M, et al. Mental health in UK Biobank – development, implementation and results from an online questionnaire completed by 157 366 participants: a reanalysis. BJPsych Open. 2020;6(2):e18. doi:10.1192/bjo.2019.100

27. Smith GD, Ebrahim S. “Mendelian randomization”: Can genetic epidemiology contribute to understanding environmental determinants of disease? Int J Epidemiol. 2003;32(1):1–22. doi:10.1093/ije/dyg070

28. Mitchell, R., Hemani, G., Dudding, T., Corbin, L., Harrison, S., Paternoster L. UK Biobank Genetic Data: MRC-IEU Quality Control, version 2 - Datasets - data.bris. DataBris. Published online 2018.

29. Gooding HC, Milliren C, McLaughlin KA, et al. Child maltreatment and blood pressure in young adulthood. Child Abuse Negl. 2014;38(11):1747–1754. doi:10.1016/j.chiabu.2014.08.019

30. Opel N, Redlich R, Repple J, et al. Childhood maltreatment moderates the influence of genetic load for obesity on reward related brain structure and function in major depression. Psychoneuroendocrinology. 2019;100(July 2018):18-26. doi:10.1016/j.psyneuen.2018.09.027

31. Liu M, Jiang Y, Wedow R, et al. Association studies of up to 1.2 million individuals yield new insights into the genetic etiology of tobacco and alcohol use. Nat Genet. 2019;51(2):237–244. doi:10.1038/s41588-018-0307-5

32. O’Neill A, Beck K, Chae D, Dyer T, He X, Lee S. The pathway from childhood maltreatment to adulthood obesity: The role of mediation by adolescent depressive symptoms and BMI. J Adolesc. 2018;67:22–30. doi:10.1016/j.adolescence.2018.05.010

33. Ruiz AL, Font SA. Role of childhood maltreatment on weight and weight-related behaviors in adulthood. Heal Psychol Off J Div Heal Psychol Am Psychol Assoc. 2020;39(11):986–996. doi:10.1037/hea0001027

34. Sokol RL, Gottfredson NC, Poti JM, et al. Sensitive Periods for the Association Between Childhood Maltreatment and BMI. Am J Prev Med. 2019;57(4):495–502. doi:10.1016/j.amepre.2019.05.016

35. Afifi TO, Mota N, MacMillan HL, Sareen J. Harsh Physical Punishment in Childhood and Adult Physical Health. Pediatrics. 2013;132(2):e333.-e340. doi:10.1542/peds.2012-4021

36. Lang J, Kerr DM, Petri-Romão P, et al. The hallmarks of childhood abuse and neglect: A systematic review. Salinas-Miranda A, ed. PLoS One. 2020;15(12):e0243639. doi:10.1371/journal.pone.0243639

37. Anda RF, Croft JB, Felitti VJ, et al. Adverse Childhood Experiences and Smoking During Adolescence and Adulthood. JAMA. 1999;282(17):1652–1658. doi:10.1001/jama.282.17.1652

38. Li L, Pinto Pereira SM, Power C. Childhood maltreatment and biomarkers for cardiometabolic disease in mid-adulthood in a prospective British birth cohort: associations and potential explanations. BMJ Open. 2019;9(3):e024079. doi:10.1136/bmjopen-2018-024079

39. van Reedt Dortland AKB, Giltay EJ, van Veen T, Zitman FG, Penninx BWJH. Personality traits and childhood trauma as correlates of metabolic risk factors: The Netherlands Study of Depression and Anxiety (NESDA). Prog Neuro-Psychopharmacology Biol Psychiatry. 2012;36(1):85–91. doi:https://doi.org/10.1016/j.pnpbp.2011.10.001

40. Lawlor DA. Biological interaction: Time to drop the term? Epidemiology. 2011;22(2):148–150. doi:10.1097/EDE.0b013e3182093298

41. Duncan LE, Keller MC. A Critical Review of the First 10 Years of Candidate Gene-by-Environment Interaction Research in Psychiatry. Am J Psychiatry. 2011;168(10):1041–1049. doi:10.1176/appi.ajp.2011.11020191

42. Choi SW, Mak TSH, O’Reilly PF. Tutorial: a guide to performing polygenic risk score analyses. Nat Protoc. 2020;15(9):2759–2772. doi:10.1038/s41596-020-0353-1

43. Tyrrell J, Zheng J, Beaumont R, et al. Genetic predictors of participation in optional components of UK Biobank. Nat Commun. 2021;12(1):886. doi:10.1038/s41467-021-21073-y

44. Moody G, Cannings-John R, Hood K, Kemp A, Robling M. Establishing the international prevalence of self-reported child maltreatment: A systematic review by maltreatment type and gender. BMC Public Health. 2018;18(1):1–15. doi:10.1186/s12889-018-6044-y

45. Haworth S, Mitchell R, Corbin L, et al. Apparent latent structure within the UK Biobank sample has implications for epidemiological analysis. Nat Commun. 2019;10(1):333. doi:10.1038/s41467-018-08219-1

46. Martin AR, Kanai M, Kamatani Y, Okada Y, Neale BM, Daly MJ. Clinical use of current polygenic risk scores may exacerbate health disparities. Nat Genet. 2019;51(4):584–591. doi:10.1038/s41588-019-0379-x

47. Mills MC, Rahal C. A scientometric review of genome-wide association studies. Commun Biol. 2019;2(1):9. doi:10.1038/s42003-018-0261-x

48. Locke AE, Kahali B, Berndt SI, et al. Genetic studies of body mass index yield new insights for obesity biology. Nature. 2015;518(7538):197–206. doi:10.1038/nature14177

49. Willer CJ, Schmidt EM, Sengupta S, et al. Discovery and refinement of loci associated with lipid levels. Nat Genet. 2013;45(11):1274–1285. doi:10.1038/ng.2797

50. Wootton RE, Richmond RC, Stuijfzand BG, et al. Evidence for causal effects of lifetime smoking on risk for depression and schizophrenia: A Mendelian randomisation study. Psychol Med. 2020;50(14):2435–2443. doi:10.1017/S0033291719002678

51. Carter AR, Gill D, Davies NM, et al. Understanding the consequences of education inequality on cardiovascular disease: Mendelian randomisation study. BMJ. 2019;365:1–12. doi:10.1136/bmj.l1855

52. Ellinor PT, Lunetta KL, Albert CM, et al. Meta-analysis identifies six new susceptibility loci for atrial fibrillation. Nat Genet. 2012;44(6):670–675. doi:10.1038/ng.2261

53. Nikpay M, Goel A, Won HH, et al. A comprehensive 1000 Genomes-based genome-wide association meta-analysis of coronary artery disease. Nat Genet. 2015;47(10):1121–1130. doi:10.1038/ng.3396

54. Scott RA, Scott LJ, Mägi R, et al. An Expanded Genome-Wide Association Study of Type 2 Diabetes in Europeans. Diabetes. 2017;66(11):2888–2902. doi:10.2337/db16-1253

55. Malik R, Chauhan G, Traylor M, et al. Multiancestry genome-wide association study of 520,000 subjects identifies 32 loci associated with stroke and stroke subtypes. Nat Genet. 2018;50(4):524–537. doi:10.1038/s41588-018-0058-3

