## Supplementary tables and methods for "Investigating childhood maltreatment as a modifier of genetic risk for cardiovascular disease in the UK Biobank"

### Supplement

*Table S1 – Questions used to determine exposure to childhood maltreatment on mental health questionnaire.*

| Question: "When I was a child…" | Type of maltreatment | Cut-off * |
|---|---|---|
| "I felt loved" | Emotional neglect | 2 or lower |
| "People in my family hit me so hard that it left me with bruises or marks" | Physical abuse | 1 or higher |
| "I felt that someone in my family hated me" | Emotional abuse | 1 or higher |
| "Someone molested me (sexually)" | Sexual abuse | 1 or higher |
| "There was someone to take me to the doctor if I needed it" | Physical neglect | 3 or lower |

*According to the following answers: 0: never true; 1: rarely true; 2: sometimes true; 3: often true; 4: very often true.

*Table S2 – Evidence supporting a causal role for the considered risk factors on cardiovascular disease risk*

| Risk factor | Study | Study design |
|---|---|---|
| Alcohol (drinks per week) | Millwood *et al,* 2019 [1] | Mendelian randomization study |
| BMI | Carter *et al,* 2019 [2]; Larsson *et al,* 2020 [3] | Mendelian randomization study; Mendelian randomization study |
| Low density lipoprotein cholesterol | Ference et al, 2017 [4]; Cheung et al, 2004 [5] | Mendelian randomization and clinical study; randomized controlled trial |
| Smoking (lifetime behaviour) | Larsson *et al,* 2020 [6] | Mendelian randomization study |
| Systolic blood pressure | Carter *et al,* 2019 [2]; Ettehad *et al,* 2016 [7] | Mendelian randomization study; systematic review including randomized controlled trial |
| Type 2 diabetes | Ahmad *et al,* 2015 [8] | Mendelian randomization study |

*Table S3 – ICD codes for the included cardiovascular diseases/risk factors.*

| Diagnosis | ICD9 | ICD10 |
|---|---|---|
| Atrial Fibrillation | 427.31 | I48 |
| Type 2 diabetes | 250.00, 250.02, 250.10, 250.12, 250.20, 250.22, 250.30, 250.32, 250.40, 250.42, 250.50, 250.52, 250.60, 250.62, 250.70, 250.72, 250.80, 250.82, 250.90, 250.92 | E11 |
| Coronary Heart Disease | 410.0-414.9 | I20-I25 |
| Stroke | 430.0-438.9 | G45, I60-I64 |

*Table S4 – Variance explained by each PGS according to the P value threshold.*

| Risk factor/disease GWAS | $P<5\times10^{-8}$ | | P<0.05 | | P<0.5 | |
|---|---|---|---|---|---|---|
| | # of SNPs | $R^2$ | # of SNPs | $R^2$ | # of SNPs | $R^2$ |
| BMI | 127 | 0.02983 | 20 542 | 0.06512 | 139 582 | 0.06868 |
| Drinks per week | 14 | 0.07834 | 72 962 | 0.08039 | 449 080 | 0.08019 |
| LDL cholesterol | 398 | 0.07026 | 13 337 | 0.02450 | 23 724 | 0.02288 |
| Lifetime smoking (sample 1) | 23 | 0.00621 | 67 741 | 0.01571 | 391 104 | 0.01606 |
| Lifetime smoking (sample 2) | 21 | 0.00641 | 66 909 | 0.01516 | 390 557 | 0.01559 |
| Systolic blood pressure (sample 1) | 126 | 0.15067 | 77 709 | 0.17562 | 373 402 | 0.17260 |
| Systolic blood pressure (sample 2) | 112 | 0.14150 | 76 557 | 0.16176 | 372 715 | 0.16011 |
| Atrial Fibrillation | 3431 | 0.09755 | 60 738 | 0.29849 | 361 969 | 0.38717 |
| Type 2 Diabetes | 18 | 0.02916 | 5 137 | 0.02422 | 134 673 | 0.02300 |
| Coronary Heart Disease | 75 | 0.06554 | 49 098 | 0.06453 | 345 040 | 0.06356 |
| Stroke | 11 | 0.03919 | 63 025 | 0.03948 | 373 240 | 0.03919 |

***Table S5 – Association of childhood maltreatment score with cardiovascular risk factors and disease.*** *Analyses were adjusted for age and sex.*

| Continuous trait | Mean difference in SD of phenotypic trait per unit increase in maltreatment score (95% CI) |
|---|---|
| BMI | 0.068 (0.062, 0.073) |
| Drinks per week | 0.002 (-0.003, 0.008) |
| LDL cholesterol | 0.003 (-0.003, 0.009) |
| Lifetime smoking | 0.101 (0.095, 0.107) |
| Systolic blood pressure | -0.011 (-0.017, -0.005) |
| **Binary trait** | **Odds ratio per unit increase in maltreatment score (95% CI)** |
| Atrial Fibrillation | 1.04 (0.99, 1.08) |
| Type 2 Diabetes | 1.19 (1.14, 1.25) |
| Coronary Heart Disease | 1.13 (1.10, 1.17) |
| Stroke | 1.06 (1.00, 1.12) |

***Table S6 – Effect modification of genetic predisposition to cardiovascular risk factors and disease by childhood maltreatment score on the additive scale at $P<5x10^{-8}$.*** *Analyses were adjusted for age, sex and 40 genetic principal components.*

| **Continuous trait** | **N** | **Additive scale** | | | |
|---|---|---|---|---|---|
| | | **Mean difference in SD of trait per SD increase of PGS (95% CI)** | **Mean difference in SD of trait per unit increase of maltreatment score (95% CI)** | **Interaction coefficient** | **P value for interaction** |
| BMI | 100659 | 0.12 (0.11, 0.13) | 0.065 (0.060, 0.071) | 0.009 (0.003, 0.014) | 0.003 |
| Drinks per week | 100529 | 0.04 (0.04, 0.05) | 0.001 (-0.004, 0.007) | 0.002 (-0.004, 0.007) | 0.598 |
| LDL cholesterol | 95809 | 0.23 (0.22, 0.23) | 0.003 (-0.002, 0.009) | 0.003 (-0.002, 0.009) | 0.238 |
| Lifetime smoking | 95845 | 0.04 (0.03, 0.04) | 0.100 (0.094, 0.105) | 0.003 (-0.003, 0.008) | 0.395 |
| Systolic blood pressure | 93001 | 0.09 (0.08, 0.10) | -0.011 (-0.016, -0.005) | -0.002 (-0.008, 0.004) | 0.490 |
| **Binary trait** | **N = 100833** | **Risk difference of trait per SD increase of PGS (95% CI)** | **Risk difference of trait per SD increase of maltreatment score (95% CI)** | **Interaction coefficient** | **P value for interaction** |
| Atrial fibrillation | Control 98735<br>Case 2098 | 0.011 (0.010, 0.012) | 0.001 (0.000, 0.002) | $4x10^{-4}$ ($-4x10^{-4}$, $1x10^{-3}$) | 0.322 |
| Type 2 diabetes | Control 99449<br>Case 1384 | 0.003 (0.003, 0.004) | 0.003 (0.002, 0.003) | $4x10^{-4}$ ($-2x10^{-4}$, $1x10^{-3}$) | 0.196 |
| Coronary heart disease | Control 97801<br>Case 3032 | 0.007 (0.006, 0.008) | 0.004 (0.003, 0.005) | $3x10^{-4}$ ($-1 x10^{-3}$, $1x10^{-3}$) | 0.611 |
| Stroke | Control 99838<br>Case 995 | $1x10^{-4}$ ($-5x10^{-4}$, $8x10^{-4}$) | 0.001 (-0.000, 0.001) | $1x10^{-5}$ ($-5x10^{-4}$, $6x10^{-4}$) | 0.973 |

***eTable 7 – Effect modification of genetic predisposition to cardiovascular risk factors and disease by childhood maltreatment score on the multiplicative scale at $P<5x10^{-8}$.*** *Analyses were adjusted for age, sex and 40 genetic principal components.*

| **Continuous trait** | **N** | **Multiplicative scale** | | | |
|---|---|---|---|---|---|
| | | **Mean difference in SD of log trait per SD increase of PGS (95% CI)** | **Mean difference in log SD of trait per unit increase of maltreatment score (95% CI)** | **Interaction coefficient** | **P value for interaction** |
| BMI | 100659 | 0.12 (0.11, 0.13) | 0.065 (0.059, 0.070) | 0.007 (0.001, 0.013) | 0.015 |
| Drinks per week | 100529 | 0.05 (0.04, 0.06) | -0.028 (-0.033, -0.022) | 0.002 (-0.004, 0.008) | 0.496 |
| LDL cholesterol | 95809 | 0.23 (0.22, 0.23) | 0.002 (-0.003, 0.008) | 0.004 (-0.002, 0.010) | 0.164 |
| Lifetime smoking | 95845 | 0.04 (0.03, 0.04) | 0.103 (0.097, 0.109) | 0.002 (-0.003, 0.008) | 0.429 |
| Systolic blood pressure | 93001 | 0.09 (0.09, 0.10) | -0.011 (-0.017, -0.006) | -0.002 (-0.007, 0.004) | 0.506 |
| **Binary trait** | **N** | **OR of trait per SD increase of PGS (95% CI)** | **OR of trait per unit increase of maltreatment score (95% CI)** | **Interaction coefficient** | **P value for interaction** |
| Atrial fibrillation | Control 98735<br>Case 2098 | 1.69 (1.62, 1.76) | 1.03 (0.98, 1.08) | 1.02 (0.98, 1.06) | 0.312 |
| Type 2 diabetes | Control 99449<br>Case 1384 | 1.29 (1.22, 1.35) | 1.20 (1.14, 1.25) | 0.99 (0.95, 1.04) | 0.743 |
| Coronary heart disease | Control 97801<br>Case 3032 | 1.28 (1.23, 1.32) | 1.14 (1.10, 1.18) | 0.99 (0.96, 1.02) | 0.615 |
| Stroke | Control 99838<br>Case 995 | 1.02 (0.95, 1.08) | 1.06 (1.00, 1.12) | 1.00 (0.94, 1.06) | 0.990 |

***Table S8 – Sex-stratified effect modification of childhood maltreatment score on genetic predisposition to cardiovascular risk factors and disease on the additive scale at P<5x10$^{-8}$.*** *Analyses were adjusted for age, sex and 40 genetic principal components. BMI=body mass index; LDL=low-density lipoprotein; PGS=polygenic score.*

| | | | Additive scale | | | | | | |
|---|---|---|---|---|---|---|---|---|---|
| **Continuous trait** | **N** | | **Sex** | **N** | **Mean difference in SD of trait per SD increase of PGS (95% CI)** | **Mean difference in SD of trait per unit increase of maltreatment score (95% CI)** | **Interaction coefficient** | **P Value for interaction** | **P value interaction by sex** |
| BMI | 100659 | | Female | 57540 | 0.13 (0.12, 0.14) | 0.067 (0.059, 0.074) | 0.007 (-0.001, 0.015) | 0.071 | 0.596 |
| | | | Male | 43119 | 0.11 (0.11, 0.12) | 0.064 (0.056, 0.072) | 0.010 (0.002, 0.019) | 0.016 | |
| Drinks per week | 100529 | | Female | 57541 | 0.04 (0.03, 0.05) | -0.002 (-0.008, 0.004) | 3E-04 (-0.006, 0.006) | 0.926 | 0.503 |
| | | | Male | 42988 | 0.05 (0.04, 0.06) | 0.005 (-0.005, 0.016) | 0.004 (-0.006, 0.015) | 0.429 | |
| LDL cholesterol | 95809 | | Female | 54698 | 0.24 (0.23, 0.25) | 0.009 (0.002, 0.016) | 0.002 (-0.005, 0.009) | 0.604 | 0.506 |
| | | | Male | 41111 | 0.21 (0.20, 0.22) | -0.002 (-0.011, 0.007) | 0.004 (-0.005, 0.013) | 0.406 | |
| Lifetime smoking | 95845 | | Female | 54768 | 0.03 (0.02, 0.04) | 0.095 (0.088, 0.102) | 0.001 (-0.006, 0.008) | 0.735 | 0.440 |
| | | | Male | 41077 | 0.04 (0.03, 0.05) | 0.107 (0.097, 0.117) | 0.006 (-0.004, 0.016) | 0.258 | |
| Systolic blood pressure | 93001 | | Female | 53143 | 0.10 (0.09, 0.11) | -0.010 (-0.018, -0.003) | -0.004 (-0.011, 0.004) | 0.320 | 0.572 |
| | | | Male | 39858 | 0.09 (0.08, 0.10) | -0.009 (-0.018, 0.000) | -2X10$^{-5}$ (-9x10$^{-3}$, 9x10$^{-3}$) | 0.972 | |
| **Binary trait** | **N = 100 833** | | **Sex**<br>**Female N= 57641**<br>**Male N = 43192** | | **Risk difference of trait per SD increase of PGS (95% CI)** | **Risk difference of trait per SD increase of maltreatment score (95% CI)** | **Interaction coefficient** | **P value for interaction** | **P value interaction by sex** |
| Atrial fibrillation | Control | 98735 | Female | | 0.007 (0.006, 0.008) | 2x10$^{-4}$ (-6x10$^{-4}$, 0.001) | 4x10$^{-4}$ (-4x10$^{-4}$, 0.001) | 0.350 | 0.432 |
| | Case | 2098 | Male | | 0.017 (0.016, 0.019) | 0.002 (0.000, 0.003) | 0.001 (-0.001, 0.003) | 0.226 | |
| Type 2 diabetes | Control | 99449 | Female | | 0.003 (0.002, 0.004) | 0.002 (0.001, 0.003) | 0.001 (0.000, 0.001) | 0.046 | 0.349 |
| | Case | 1384 | Male | | 0.004 (0.003, 0.005) | 0.004 (0.003, 0.005) | 6x10-5 (-0.001, 0.001) | 0.926 | |
| Coronary heart disease | Control | 97801 | Female | | 0.004 (0.003, 0.005) | 0.003 (0.002, 0.004) | -3x10$^{-4}$ (-1x10-3, 1x10-3) | 0.527 | 0.065 |
| | Case | 3032 | Male | | 0.011 (0.009, 0.013) | 0.004 (0.002, 0.006) | 0.002 (-4x10$^{-4}$, 0.004) | 0.116 | |
| Stroke | Control | 99838 | Female | | 2x10$^{-4}$ (-5x10$^{-4}$, 9x10$^{-4}$) | 0.001 (0.000, 0.001) | 5x10$^{-4}$ (-1x10$^{-4}$, 1x10$^{-3}$) | 0.106 | 0.020 |
| | Case | 995 | Male | | 1x10$^{-4}$ (-1x10$^{-3}$, 1x10$^{-3}$) | 2x10$^{-4}$ (-9x10$^{-4}$, 1x10$^{-3}$) | -1x10$^{-4}$ (-2x10$^{-3}$, 2x10$^{-4}$) | 0.118 | |

*Table S9 – **Sex-stratified effect modification of childhood maltreatment score on genetic predisposition to cardiovascular risk factors and disease on the multiplicative scale at $P<5x10^{-8}$.** Analyses were adjusted for age, sex and 40 genetic principal components. BMI=body mass index; LDL=low-density lipoprotein; PGS=polygenic score.*

| Continuous trait | N | | Sex | N | Multiplicative scale: Mean difference in SD of log trait per SD increase of PGS (95% CI) | Mean difference in log SD of trait per unit increase of maltreatment score (95% CI) | Interaction coefficient | P value for Interaction | P value interaction by sex |
|---|---|---|---|---|---|---|---|---|---|
| BMI | 100659 | | Female | 57540 | 0.13 (0.12, 0.14) | 0.066 (0.059, 0.074) | 0.006 (-0.002, 0.013) | 0.145 | 0.635 |
| | | | Male | 43119 | 0.11 (0.11, 0.12) | 0.062 (0.053, 0.070) | 0.009 (3x10-4, 0.017) | 0.042 | |
| Drinks per week | 100529 | | Female | 57541 | 0.05 (0.05, 0.06) | -0.028 (-0.035, -0.021) | 0.001 (-0.006, 0.008) | 0.839 | 0.616 |
| | | | Male | 42988 | 0.04 (0.03, 0.05) | -0.028 (-0.037, -0.019) | 0.004 (-0.005, 0.013) | 0.402 | |
| LDL cholesterol | 95809 | | Female | 54698 | 0.24 (0.23, 0.25) | 0.008 (0.001, 0.015) | 0.003 (-0.004, 0.010) | 0.465 | 0.544 |
| | | | Male | 41111 | 0.21 (0.20, 0.21) | -0.003 (-0.013, 0.006) | 0.004 (-0.005, 0.014) | 0.379 | |
| Lifetime smoking | 95845 | | Female | 54768 | 0.03 (0.02, 0.04) | 0.099 (0.092, 0.106) | 0.002 (-0.005, 0.009) | 0.620 | 0.672 |
| | | | Male | 41077 | 0.05 (0.04, 0.06) | 0.108 (0.098, 0.119) | 0.004 (-0.006, 0.015) | 0.396 | |
| Systolic blood pressure | 93001 | | Female | 53143 | 0.10 (0.09, 0.11) | -0.011 (-0.018, -0.004) | -0.004 (-0.011, 0.003) | 0.280 | 0.477 |
| | | | Male | 39858 | 0.09 (0.08, 0.09) | -0.009 (-0.018, -0.001) | 4x10-4 (-0.008, 0.009) | 0.934 | |
| **Binary trait** | **N = 100 833** | | **Sex Female N= 57641 Male N = 43192** | | **OR of trait per SD increase of PGS (95% CI)** | **OR of trait per unit increase of maltreatment score (95% CI)** | **Interaction coefficient** | **P value for Interaction** | **P value interaction by sex** |
| Atrial fibrillation | Control | 98735 | Female | | 1.61 (1.50, 1.72) | 1.00 (0.93, 1.07) | 1.03 (0.97, 1.10) | 0.291 | 0.683 |
| | Case | 2098 | Male | | 1.74 (1.65, 1.84) | 1.05 (0.99, 1.12) | 1.01 (0.96, 1.07) | 0.588 | |
| Type 2 diabetes | Control | 99449 | Female | | 1.34 (1.24, 1.46) | 1.18 (1.10, 1.26) | 1.01 (0.95, 1.08) | 0.651 | 0.324 |
| | Case | 1384 | Male | | 1.25 (1.17, 1.33) | 1.22 (1.14, 1.29) | 0.97 (0.91, 1.03) | 0.341 | |
| Coronary heart disease | Control | 97801 | Female | | 1.23 (1.16, 1.31) | 1.20 (1.14, 1.26) | 0.96 (0.92, 1.01) | 0.127 | 0.088 |
| | Case | 3032 | Male | | 1.30 (1.24, 1.36) | 1.09 (1.04, 1.14) | 1.02 (0.97, 1.07) | 0.401 | |
| Stroke | Control | 99838 | Female | | 1.03 (0.93, 1.13) | 1.09 (1.01, 1.19) | 1.06 (0.98, 1.15) | 0.136 | 0.029 |
| | Case | 995 | Male | | 1.01 (0.93, 1.10) | 1.01 (0.93, 1.10) | 0.94 (0.86, 1.02) | 0.123 | |

***Table S10 – Effect modification of genetic predisposition to cardiovascular risk factors and disease by childhood maltreatment score on the additive and multiplicative scale at P<0.05.*** *Analyses were adjusted for age, sex and 40 genetic principal components. BMI=body mass index; LDL=low-density lipoprotein; PGS=polygenic score.*

| | **P<0.05** | | | | | | | |
|---|---|---|---|---|---|---|---|---|
| | **Additive scale** | | | | **Multiplicative scale** | | | |
| **Continuous Trait** | **Mean difference in SD of trait per SD increase of PGS (95% CI)** | **Mean difference in SD of trait per unit increase of maltreatment score (95% CI)** | **Interaction coefficient** | **P value of interaction** | **Mean difference in SD of log trait per SD increase of PGS (95% CI)** | **Mean difference in log SD of trait per unit increase of maltreatment score (95% CI)** | **Interaction coefficient** | **P value of interaction** |
| BMI | 0.23 (0.22, 0.23) | 0.061 (0.056, 0.067) | 0.016 (0.010, 0.021) | $2x10^{-8}$ | 0.23 (0.22, 0.23) | 0.060 (0.055, 0.066) | 0.013 (0.008, 0.019) | $4x10^{-6}$ |
| Drinks per week | 0.06 (0.06, 0.07) | 0.001 (-0.005, 0.007) | -0.002 (-0.007, 0.004) | 0.465 | 0.06 (0.06, 0.07) | -0.028 (-0.033, -0.022) | -0.002 (-0.008, 0.004) | 0.480 |
| LDL cholesterol | 0.08 (0.07, 0.09) | 0.003 (-0.003, 0.009) | $3x10^{-4}$ (-$6x10^{-3}$, $6x10^{-3}$) | 0.917 | 0.08 (0.07, 0.09) | 0.002 (-0.004, 0.008) | $2x10^{-4}$ (-$6x10^{-3}$, $6x10^{-3}$) | 0.941 |
| Lifetime smoking | 0.12 (0.11, 0.13) | 0.206 (0.143, 0.269) | 0.017 (0.009, 0.024) | $7x10^{-6}$ | 0.12 (0.12, 0.13) | 0.215 (0.152, 0.278) | 0.014 (0.007, 0.022) | $1x10^{-4}$ |
| Systolic blood pressure | 0.21 (0.20, 0.22) | 0.043 (-0.017, 0.101) | -$9x10^{-5}$ (-$7x10^{-3}$, $6x10^{-3}$) | 0.978 | 0.21 (0.21, 0.22) | 0.037 (-0.022, 0.095) | 0.001 (-0.006, 0.007) | 0.223 |
| **Binary Trait** | **Risk difference of trait per SD increase of PGS (95% CI)** | **Risk difference of trait per SD increase of maltreatment score (95% CI)** | **Interaction coefficient** | **P value of interaction** | **OR of trait per SD increase of PGS (95% CI)** | **OR of trait per unit increase of maltreatment score (95% CI)** | **Interaction coefficient** | **P value of interaction** |
| Atrial fibrillation | 0.032 (0.031, 0.033) | 0.001 (0.000, 0.001) | -$6x10^{-4}$ (-$1x10^{-4}$, $2x10^{-4}$) | 0.122 | 4.56 (4.35, 4.79) | 1.06 (0.98, 1.14) | 0.98 (0.93, 1.02) | 0.365 |
| Type 2 diabetes | 0.002 (0.001, 0.002) | 0.003 (0.002, 0.003) | $2x10^{-4}$ (-$5x10^{-4}$, $9x10^{-4}$) | 0.575 | 1.12 (1.06, 1.18) | 1.20 (1.14, 1.25) | 1.00 (0.95, 1.04) | 0.867 |
| Coronary heart disease | 0.006 (0.005, 0.007) | 0.004 (0.003, 0.004) | -$3x10^{-5}$ (-$1x10^{-3}$, $1x10^{-3}$) | 0.954 | 1.25 (1.20, 1.30) | 1.13 (1.10, 1.17) | 0.98 (0.95, 1.02) | 0.367 |
| Stroke | 0.001 (0.000, 0.001) | 0.001 (0.000, 0.001) | -1x10-5 (-6x10-4, $6x10^{-4}$) | 0.960 | 1.06 (1.00, 1.13) | 1.06 (0.99, 1.12) | 1.00 (0.94, 1.06) | 0.953 |

***Table S11 – Effect modification of genetic predisposition to cardiovascular risk factors and disease by childhood maltreatment score on the additive and multiplicative scale at P<0.5.*** *Analyses were adjusted for age, sex and 40 genetic principal components. BMI=body mass index; LDL=low-density lipoprotein; PGS=polygenic score.*

| | **P<0.5** | | | | | | | |
|---|---|---|---|---|---|---|---|---|
| | **Additive scale** | | | | **Multiplicative scale** | | | |
| **Continuous Trait** | **Mean difference in SD of trait per SD increase of PGS (95% CI)** | **Mean difference in SD of trait per unit increase of maltreatment score (95% CI)** | **Interaction coefficient** | **P value of interaction** | **Mean difference in SD of log trait per SD increase of PGS (95% CI)** | **Mean difference in log SD of trait per unit increase of maltreatment score (95% CI)** | **Interaction coefficient** | **P value of interaction** |
| BMI | 0.23 (0.23, 0.24) | 0.061 (0.055, 0.066) | 0.015 (0.009, 0.020) | $1 \times 10^{-7}$ | 0.24 (0.23, 0.24) | 0.060 (0.054, 0.066) | 0.011 (0.007, 0.017) | $2 \times 10^{-5}$ |
| Drinks per week | 0.06 (0.06, 0.07) | 0.001 (-0.005, 0.007) | -0.002 (-0.008, 0.003) | 0.467 | 0.06 (0.06, 0.07) | -0.028 (-0.033, -0.022) | -0.002 (-0.008, 0.003) | 0.397 |
| LDL cholesterol | 0.07 (0.06, 0.08) | 0.003 (-0.003, 0.009) | 0.003 (-0.003, 0.009) | 0.276 | 0.07 (0.06, 0.07) | 0.002 (-0.004, 0.008) | 0.003 (-0.003, 0.008) | 0.377 |
| Lifetime smoking | 0.14 (0.13, 0.14) | 0.201 (0.138, 0.264) | 0.022 (0.014, 0.030) | $3 \times 10^{-8}$ | 0.14 (0.13, 0.15) | 0.210 (0.147, 0.273) | 0.020 (0.012, 0.028) | $9 \times 10^{-7}$ |
| Systolic blood pressure | 0.22 (0.21, 0.23) | 0.036 (-0.023, 0.096) | 0.002 (-0.005, 0.009) | 0.574 | 0.22 (0.22, 0.23) | 0.031 (-0.028, 0.090) | 0.003 (-0.004, 0.010) | 0.413 |
| **Binary Trait** | **Risk difference of trait per SD increase of PGS (95% CI)** | **Risk difference of trait per SD increase of maltreatment score (95% CI)** | **Interaction coefficient** | **P value of interaction** | **OR of trait per SD increase of PGS (95% CI)** | **OR of trait per unit increase of maltreatment score (95% CI)** | **Interaction coefficient** | **P value of interaction** |
| Atrial fibrillation | 0.040 (0.039, 0.041) | 0.001 (-0.000, 0.001) | -0.001 (-0.002, 0.000) | 0.066 | 5.37 (5.11, 5.64) | 1.06 (0.97, 1.15) | 0.98 (0.94, 1.03) | 0.432 |
| Type 2 diabetes | $2 \times 10^{-4}$ ($-5 \times 10^{-4}$, $9 \times 10^{-4}$) | 0.003 (0.002, 0.003) | $2 \times 10^{-4}$ ($-5 \times 10^{-4}$, $9 \times 10^{-4}$) | 0.535 | 1.02 (0.96, 1.07) | 1.20 (1.14, 1.25) | 1.01 (0.97, 1.06) | 0.615 |
| Coronary heart disease | 0.006 (0.005, 0.007) | 0.004 (0.003, 0.005) | $-1 \times 10^{-4}$ ($-1 \times 10^{-3}$, $8 \times 10^{-4}$) | 0.782 | 1.22 (1.18, 1.27) | 1.14 (1.10, 1.17) | 0.98 (0.95, 1.02) | 0.331 |
| Stroke | $1 \times 10^{-4}$ ($-5 \times 10^{-4}$, $8 \times 10^{-4}$) | 0.001 (0.000, 0.001) | $4 \times 10^{-5}$ ($-5 \times 10^{-4}$, $6 \times 10^{-4}$) | 0.901 | 1.02 (0.95, 1.08) | 1.06 (1.00, 1.12) | 1.01 (0.95, 1.07) | 0.865 |

***Table S12 – Effect modification of genetic predisposition to cardiovascular risk factors and disease by childhood maltreatment score on the additive and multiplicative scale at P<5x10$^{-8}$, adjusted for covariate interactions.*** *Interactions considered were those between main confounders (age, sex, and genetic principal components) and the polygenic scores or maltreatment scores. Analyses were also adjusted for age, sex and 40 genetic principal components. BMI=body mass index; LDL=low-density lipoprotein; PGS=polygenic score.*

| | **Adjusted for covariate interactions, P<5x10$^{-8}$** | | | | | | | |
|---|---|---|---|---|---|---|---|---|
| | **Additive scale** | | | | **Multiplicative scale** | | | |
| **Continuous Trait** | **Mean difference in SD of trait per SD increase of PGS (95% CI)** | **Mean difference in SD of trait per unit increase of maltreatment score (95% CI)** | **Interaction coefficient** | **P value of interaction** | **Mean difference in SD of log trait per SD increase of PGS (95% CI)** | **Mean difference in log SD of trait per unit increase of maltreatment score (95% CI)** | **Interaction coefficient** | **P value of interaction** |
| BMI | 0.12 (0.11, 0.13) | 0.099 (0.038, 0.160) | 0.007 (0.001, 0.012) | 0.015 | 0.12 (0.11, 0.13) | 0.094 (0.033, 0.155) | 0.006 (-0.000, 0.011) | 0.054 |
| Drinks per week | 0.04 (0.04, 0.05) | -0.004 (-0.063, 0.056) | 0.002 (-0.004, 0.007) | 0.543 | 0.05 (0.04, 0.06) | -0.068 (-0.129, -0.008) | 0.002 (-0.004, 0.007) | 0.598 |
| LDL cholesterol | 0.23 (0.22, 0.23) | 0.011 (-0.050, 0.072) | 0.002 (-0.004, 0.008) | 0.476 | 0.23 (0.22, 0.23) | 0.014 (-0.047, 0.076) | 0.002 (-0.004, 0.008) | 0.452 |
| Lifetime smoking | 0.04 (0.03, 0.04) | 0.215 (0.152, 0.279) | 0.002 (-0.003, 0.008) | 0.463 | 0.04 (0.03, 0.04) | 0.224 (0.161, 0.287) | 0.002 (-0.004, 0.008) | 0.515 |
| Systolic blood pressure | 0.09 (0.08, 0.10) | 0.039 (-0.021, 0.099) | -0.002 (-0.008, 0.004) | 0.395 | 0.09 (0.09, 0.10) | 0.034 (-0.026, 0.093) | -0.003 (-0.008, 0.003) | 0.366 |
| **Binary Trait** | **Risk difference of trait per SD increase of PGS (95% CI)** | **Risk difference of trait per SD increase of maltreatment score (95% CI)** | **Interaction coefficient** | **P value of interaction** | **OR of trait per SD increase of PGS (95% CI)** | **OR of trait per unit increase of maltreatment score (95% CI)** | **Interaction coefficient** | **P value of interaction** |
| Atrial fibrillation | 0.011 (0.010, 0.012) | -0.002 (-0.011, 0.006) | 0.001 (0.000, 0.002) | 0.022 | 1.69 (1.62, 1.76) | 0.91 (0.54, 1.56) | 1.02 (0.98, 1.06) | 0.284 |
| Type 2 diabetes | 0.003 (0.003, 0.004) | 0.002 (-0.005, 0.009) | 4 x10$^{-4}$ (-2x10$^{-4}$, 1x10$^{-4}$) | 0.173 | 1.29 (1.22, 1.35) | 1.44 (0.88, 2.36) | 0.99 (0.95, 1.03) | 0.656 |
| Coronary heart disease | 0.007 (0.006, 0.008) | -0.002 (-0.012, 0.009) | 0.001 (-0.000, 0.002) | 0.261 | 1.28 (1.23, 1.32) | 1.58 (1.08, 2.31) | 0.99 (0.96, 1.03) | 0.657 |
| Stroke | 1x10$^{-4}$ (-5x10$^{-4}$, 8x10$^{-4}$) | -0.003 (-0.009, 0.003) | 5x10$^{-5}$ (-5x10$^{-4}$, 6x10$^{-4}$) | 0.856 | 1.02 (0.95, 1.08) | 0.86 (0.43, 1.72) | 1.00 (0.95, 1.06) | 0.887 |

***Table S13 – Effect modification of genetic predisposition to cardiovascular risk factors and disease by exposure to childhood maltreatment on the additive and multiplicative scale at P<5x10-8, adjusted for childhood SEP.*** *Proxies of childhood SEP utilised were number of siblings and maternal smoking. Analyses were also adjusted for age, sex and 40 genetic principal components. BMI=body mass index; LDL=low-density lipoprotein; PGS=polygenic score.*

| | **Adjusted for childhood SEP, P<5x10$^{-8}$** | | | | | | | |
|---|---|---|---|---|---|---|---|---|
| | **Additive scale** | | | | **Multiplicative scale** | | | |
| **Continuous Trait** | **Mean difference in SD of trait per SD increase of PGS (95% CI)** | **Mean difference in SD of trait per unit increase of maltreatment score (95% CI)** | **Interaction coefficient** | **P value of interaction** | **Mean difference in SD of log trait per SD increase of PGS (95% CI)** | **Mean difference in log SD of trait per unit increase of maltreatment score (95% CI)** | **Interaction coefficient** | **P value of interaction** |
| BMI | 0.12 (0.12, 0.13) | 0.057 (0.051, 0.063) | 0.010 (0.004, 0.016) | 0.002 | 0.12 (0.12, 0.13) | 0.056 (0.050, 0.062) | 0.009 (0.003, 0.015) | 0.006 |
| Drinks per week | 0.04 (0.04, 0.05) | 0.002 (-0.004, 0.008) | 4x10$^{-4}$ (-6x10$^{-3}$, 6x10$^{-3}$) | 0.905 | 0.05 (0.04, 0.05) | -0.025 (-0.031, -0.019) | 0.001 (-0.005, 0.007) | 0.807 |
| LDL cholesterol | 0.23 (0.22, 0.24) | 2x10$^{-4}$ (-6x10$^{-4}$, 6x10$^{-4}$) | 0.004 (-0.002, 0.010) | 0.160 | 0.23 (0.22, 0.23) | -0.001 (-0.007, 0.005) | 0.005 (-0.001, 0.011) | 0.091 |
| Lifetime smoking | 0.04 (0.03, 0.04) | 0.087 (0.081, 0.093) | 0.003 (-0.003, 0.010) | 0.269 | 0.04 (0.03, 0.04) | 0.090 (0.084, 0.096) | 0.004 (-0.003, 0.010) | 0.249 |
| Systolic blood pressure | 0.09 (0.09, 0.10) | -0.010 (-0.016, -0.004) | -0.001 (-0.007, 0.005) | 0.776 | 0.09 (0.09, 0.10) | -0.011 (-0.017, -0.005) | -0.001 (-0.007, 0.005) | 0.795 |
| **Binary Trait** | **Risk difference of trait per SD increase of PGS (95% CI)** | **Risk difference of trait per SD increase of maltreatment score (95% CI)** | **Interaction coefficient** | **P value of interaction** | **OR of trait per SD increase of PGS (95% CI)** | **OR of trait per unit increase of maltreatment score (95% CI)** | **Interaction coefficient** | **P value of interaction** |
| Atrial fibrillation | 0.011 (0.010, 0.012) | 0.001 (-0.000, 0.001) | 0.001 (-0.000, 0.001) | 0.227 | 1.72 (1.64, 1.79) | 1.01 (0.96, 1.06) | 1.03 (0.99, 1.07) | 0.183 |
| Type 2 diabetes | 0.003 (0.003, 0.004) | 0.002 (0.002, 0.003) | 0.001 (-0.000, 0.001) | 0.082 | 1.29 (1.22, 1.36) | 1.17 (1.12, 1.23) | 1.00 (0.96, 1.05) | 0.865 |
| Coronary heart disease | 0.007 (0.005, 0.008) | 0.003 (0.002, 0.004) | 0.001 (-0.001, 0.002) | 0.320 | 1.27 (1.22, 1.32) | 1.12 (1.08, 1.17) | 1.00 (0.97, 1.04) | 0.963 |
| Stroke | 6x10$^{-5}$ (-6x10$^{-4}$, 7x10$^{-4}$) | 5x10$^{-4}$ (-1x10$^{-4}$, 1x10$^{-3}$) | 2x10$^{-5}$ (-6 x10$^{-4}$, 6x10$^{-4}$) | 0.956 | 1.01 (0.94, 1.08) | 1.05 (0.99, 1.12) | 1.00 (0.94, 1.07) | 0.970 |

### Supplementary Methods 1: Polygenic scores (PGS)

*Derivation of PGSs*

Summary statistics were downloaded from MR-Base [9] using the R statistical package, including rs IDs, effect estimates, effect alleles and relevant study information. The 1000 Genomes Project was used to find proxy SNPs in LD ($R^2$ above 0.8) with SNPs not found in UK Biobank. The SNPs from MR-Base were harmonised with the SNPs from UK Biobank, aligning the effect estimates and alleles. SNPs were clumped using the European subsample of the 1000 genomes project, with $R^2 < 0.001$ and a window of 10,000 kb. PGS were calculated by multiplying the number of effect alleles for each participant in UK Biobank by the effect estimate of the SNP, then summing across all SNPs associated with each trait.

Suitable summary statistics were not available for SBP and lifetime smoking, thus a split sample GWAS of the UK Biobank was performed to generate PGSs. Briefly, the participants of the study were randomly allocated to one of two samples and a GWAS was performed on each of the samples. Summary statistics of each sample were then used to calculate PGSs for the other sample, avoiding sample overlap.[10] This method has been described in detail previously.[2]

*BMI*

BMI ($kg/m^2$) was calculated using the height and weight measurements collected at baseline.

We used summary statistics from the Genetic Investigation of Anthropometric Traits (GIANT) Consortium BMI GWAS on individuals of European ancestry (n=339 224),[11] the most recent BMI GWAS without UK Biobank participants to avoid sample overlap.

*Alcohol consumption (drinks per week)*

Alcohol consumption was self-reported by participants at baseline at the assessment centres. Participants were asked to report their current drinking behaviours with the options daily or almost daily, three or four times a week, once or twice a week, one to three times a month, special occasions only, never or prefer not to say. Participants with an intake of once or twice weekly and over were asked to report their weekly intake of a number of alcoholic beverages; red wine, white wine, champagne, beer, cider, spirits, fortified wine. These were summed to create a variable reflecting the number of drinks per week, in which those which reported to drink one to three times a month or less were assumed to have an intake of 0 drinks per week. This variable has been previously described elsewhere.[12]

Summary statistics from the GWAS and Sequencing Consortium of Alcohol and Nicotine use (GSCAN) GWAS of drinks per week, as previously described,[13] were employed to generate the PGSs. UK Biobank participants were excluded to avoid sample overlap.

*LDL cholesterol*

LDL cholesterol was measured from plasma samples collected from participants by enzymatic protective selection analysis on a Beckman Coulter AU5800. Measurements were excluded by UK Biobank if they did not pass the UK Biobank's quality control checks after an inadvertent dilution of samples.[14]

Summary statistics were taken from the Global Lipids Genetics Consortium LDL cholesterol GWAS of individuals of European ancestry (n=188,577).[15]

*Lifetime smoking*

Smoking behaviour was self-reported by participants at baseline at the assessment centres. Participants were asked about smoking status (current, former or never), age of initiation, and cigarettes smoked per day. A variable reflecting lifetime smoking behaviour was generated by combining the smoking measures into an index and a half-life constant reflecting the exponential decrease in effect of smoking at a given time on health. A detailed overview of the construction of this variable can be found elsewhere. [16]

For the GWAS, estimates from the statistical analyses using the two samples were meta-analysed by using the *metan* command on Stata.

*Systolic blood pressure*

Participant systolic blood pressure measurements were taken at baseline while attending assessment centres using an Omron HEM-7105IT blood pressure monitor. The mean of two resting blood pressure measurements was used.

For the GWAS summary statistics, estimates from the statistical analyses using the two samples were meta-analysed by using the *metan* command on Stata.

*Atrial Fibrillation*

Atrial fibrillation cases were determined through mortality data from the UK Biobank and linked hospital episode statistics and Scottish morbidity and mortality records, referred to jointly as hospital inpatient records. Cases were identified according to the specified ICD9 and ICD10 disease code as shown in Table S3. Date of diagnosis and date of attendance of assessment centre were compared to distinguish prevalent from incident cases.

Summary statistics were obtained from an Atrial Fibrillation GWAS of individuals of European ancestry (n=59 133, of which 6 707 with atrial fibrillation).[17]

*Type 2 Diabetes*

T2DM cases were determined through mortality data from the UK Biobank and linked hospital inpatient records. Date of diagnosis and date of attendance of assessment centre were compared to distinguish prevalent from incident cases. Given the relatively low proportion of hospitalisation episodes attributed to T2DM alone, self-reported cases to UK Biobank nurses at an assessment centre were included. Cases reported at baseline were identified as prevalent if reported at baseline and as incident if reported at a follow-up visit to the assessment centre, though only a proportion of participants attend these. These self-reported variables do not distinguish between type 1 and type 2 diabetes, yet most of these cases are expected to be of the latter.

Summary statistics were taken from the DIAbetes Genetics Replication And Meta-analysis (DIAGRAM) Consortium GWAS of individuals of European ancestry (n=159,234 of which 26,676 with T2DM).[18]

*Coronary Heart Disease*

CHD cases were determined through mortality data from the UK Biobank and linked hospital inpatient records. Cases were identified according to the specified ICD9 and ICD10 disease codes as shown in Table S3, spanning subtypes such as acute myocardial infarction and angina pectoris. Date of diagnosis and date of attendance of assessment centre were compared to distinguish prevalent from incident cases.

Summary statistics were taken from the most recent CHD GWAS without UK Biobank participants. The GWAS included individuals of predominantly (77%) European ancestry (n=184,305, of which 60,801 with CHD). [19]

*Stroke*

Stroke cases were determined through mortality data from the UK Biobank and linked hospital inpatient records. Cases were identified according to the specified ICD9 and ICD10 disease code as shown in Table S3. Date of diagnosis and date of attendance of assessment centre were compared to distinguish prevalent from incident cases.

Summary statistics were obtained from the MEGASTROKE Consortium GWAS including individuals European ancestry (n=446,696, of which cases = 40,585).[20]

**References**

1. Millwood IY, Walters RG, Mei XW, et al. Conventional and genetic evidence on alcohol and vascular disease aetiology: a prospective study of 500 000 men and women in China. *Lancet*. 2019;393(10183):1831-1842. doi:10.1016/S0140-6736(18)31772-0

2. Carter AR, Gill D, Davies NM, et al. Understanding the consequences of education inequality on cardiovascular disease: Mendelian randomisation study. *BMJ*. 2019;365:1-12. doi:10.1136/bmj.l1855

3. Larsson SC, Bäck M, Rees JMB, Mason AM, Burgess S. Body mass index and body composition in relation to 14 cardiovascular conditions in UK Biobank: a Mendelian randomization study.

*Eur Heart J*. 2020;41(2):221-226. doi:10.1093/eurheartj/ehz388

4. Ference BA, Ginsberg HN, Graham I, et al. Low-density lipoproteins cause atherosclerotic cardiovascular disease. 1. Evidence from genetic, epidemiologic, and clinical studies. A consensus statement from the European Atherosclerosis Society Consensus Panel. *Eur Heart J*. 2017;38(32):2459-2472. doi:10.1093/eurheartj/ehx144

5. Cheung BMY, Lauder IJ, Lau C-P, Kumana CR. Meta-analysis of large randomized controlled trials to evaluate the impact of statins on cardiovascular outcomes. *Br J Clin Pharmacol*. 2004;57(5):640-651. doi:10.1111/j.1365-2125.2003.02060.x

6. Larsson SC, Mason AM, Bäck M, et al. Genetic predisposition to smoking in relation to 14 cardiovascular diseases. *Eur Heart J*. 2020;41(35):3304-3310. doi:10.1093/eurheartj/ehaa193

7. Ettehad D, Emdin CA, Kiran A, et al. Blood pressure lowering for prevention of cardiovascular disease and death: a systematic review and meta-analysis. *Lancet*. 2016;387(10022):957-967. doi:10.1016/S0140-6736(15)01225-8

8. Ahmad OS, Morris JA, Mujammami M, et al. A Mendelian randomization study of the effect of type-2 diabetes on coronary heart disease. *Nat Commun*. 2015;6:7060. doi:10.1038/ncomms8060

9. Hemani G, Zheng J, Elsworth B, et al. The MR-Base platform supports systematic causal inference across the human phenome. Loos R, ed. *Elife*. 2018;7:e34408. doi:10.7554/eLife.34408

10. Choi SW, Mak TSH, O'Reilly PF. Tutorial: a guide to performing polygenic risk score analyses. *Nat Protoc*. 2020;15(9):2759-2772. doi:10.1038/s41596-020-0353-1

11. Locke AE, Kahali B, Berndt SI, et al. Genetic studies of body mass index yield new insights for obesity biology. *Nature*. 2015;518(7538):197-206. doi:10.1038/nature14177

12. Howe LJ, Lawson DJ, Davies NM, et al. Genetic evidence for assortative mating on alcohol consumption in the UK Biobank. *Nat Commun*. 2019;10(1). doi:10.1038/s41467-019-12424-x

13. Liu M, Jiang Y, Wedow R, et al. Association studies of up to 1.2 million individuals yield new insights into the genetic etiology of tobacco and alcohol use. *Nat Genet*. 2019;51(2):237-244. doi:10.1038/s41588-018-0307-5

14. UK Biobank. Biomarker assay quality procedures: approaches used to minimise systematic and random errors (and the wider epidemiological implications). 2019;(April):25.

15. Willer CJ, Schmidt EM, Sengupta S, et al. Discovery and refinement of loci associated with lipid levels. *Nat Genet*. 2013;45(11):1274-1285. doi:10.1038/ng.2797

16. Wootton RE, Richmond RC, Stuijfzand BG, et al. Evidence for causal effects of lifetime smoking on risk for depression and schizophrenia: A Mendelian randomisation study. *Psychol Med*. 2020;50(14):2435-2443. doi:10.1017/S0033291719002678

17. Ellinor PT, Lunetta KL, Albert CM, et al. Meta-analysis identifies six new susceptibility loci for atrial fibrillation. *Nat Genet*. 2012;44(6):670-675. doi:10.1038/ng.2261

18. Scott RA, Scott LJ, Mägi R, et al. An Expanded Genome-Wide Association Study of Type 2 Diabetes in Europeans. *Diabetes*. 2017;66(11):2888-2902. doi:10.2337/db16-1253

19. Nikpay M, Goel A, Won HH, et al. A comprehensive 1000 Genomes-based genome-wide association meta-analysis of coronary artery disease. *Nat Genet*. 2015;47(10):1121-1130. doi:10.1038/ng.3396

20. Malik R, Chauhan G, Traylor M, et al. Multiancestry genome-wide association study of 520,000 subjects identifies 32 loci associated with stroke and stroke subtypes. *Nat Genet*.

2018;50(4):524-537. doi:10.1038/s41588-018-0058-3
